# Rare Genomic Copy Number Variants Implicate New Candidate Genes for Bicuspid Aortic Valve

**DOI:** 10.1101/2023.10.23.23297397

**Authors:** Steven G. Carlisle, Hasan Albasha, Hector Michelena, Anna Sabate-Rotes, Lisa Bianco, Julie De Backer, Laura Muiño Mosquera, Anji T. Yetman, Malenka M Bissell, Maria Grazia Andreassi, Ilenia Foffa, Dawn S. Hui, Anthony Caffarelli, Yuli Y. Kim, Dong-Chuan Guo, Rodolfo Citro, Margot De Marco, Justin T. Tretter, Kim L. McBride, EBAV Investigators, BAVCon Investigators, Dianna M. Milewicz, Simon C. Body, Siddharth K. Prakash

## Abstract

Bicuspid aortic valve (BAV), the most common congenital heart defect, is a major cause of aortic valve disease requiring valve interventions and thoracic aortic aneurysms predisposing to acute aortic dissections. The spectrum of BAV ranges from early onset valve and aortic complications (EBAV) to sporadic late onset disease. Rare genomic copy number variants (CNVs) have previously been implicated in the development of BAV and thoracic aortic aneurysms. We determined the frequency and gene content of rare CNVs in EBAV probands (n = 272) using genome-wide SNP microarray analysis and three complementary CNV detection algorithms (cnvPartition, PennCNV, and QuantiSNP). Unselected control genotypes from the Database of Genotypes and Phenotypes were analyzed using identical methods. We filtered the data to select large genic CNVs that were detected by multiple algorithms. Findings were replicated in cohorts with late onset sporadic disease (n = 5040). We identified 34 large and rare (< 1:1000 in controls) CNVs in EBAV probands. The burden of CNVs intersecting with genes known to cause BAV when mutated was increased in case-control analysis. CNVs intersecting with *GATA4* and *DSCAM* were enriched in cases, recurrent in other datasets, and segregated with disease in families. In total, we identified potentially pathogenic CNVs in 8% of EBAV cases, implicating alterations of candidate genes at these loci in the pathogenesis of BAV.

**Author Summary:** Bicuspid aortic valve (BAV) is the most common form of congenital heart disease and can lead to long-term complications such as aortic stenosis, aortic regurgitation, or thoracic aortic aneurysms. Most BAV-related complications arise in late adulthood, but 10-15% of individuals with BAV develop early onset complications before age 30. Copy number variants (CNVs) are genomic structural variations that have been previously implicated in some types of congenital heart disease, including BAV. Here we demonstrate that individuals with early onset complications of BAV are enriched for specific rare CNVs compared to individuals with late-onset BAV disease. We also describe novel CNVs involving *DSCAM*, a gene on chromosome 21 that has not previously been associated with the development of BAV. These results may lead to improved risk stratification and targeted therapies for BAV patients.

## Introduction

Copy number variants (CNVs) have been implicated as causes or modifiers of many human diseases [1]. Specifically, large genomic CNVs are significantly enriched in cohorts with developmental delay or congenital abnormalities, and the severity of phenotypes has been correlated with the burden of rare CNVs [2]. These observations show that large, rare, *de novo* CNVs are likely to be pathogenic and can exert clinically relevant effects on disease pathogenesis [3-4].

Congenital heart disease (CHD) has a worldwide prevalence of 8.2 per 1000 live births [5]. CNVs have been implicated in both syndromic and non-syndromic forms of CHD [6-10]. The pathogenicity and penetrance of CNVs was initially established for clinical syndromes such as velocardiofacial syndrome, Turner syndrome, or Williams–Beuren syndrome, which involve chromosomal or megabase scale duplications or deletions, but has since been expanded to include additional CHD subtypes [10]. CNVs contribute to 10% of all CHD cases and up to 25% of cases with extracardiac anomalies or other syndromic features [11]. The role of pathogenic CNVs affecting genes that are known to cause CHD when mutated, such as *GATA4* and *TBX1*, has been established [12]. Furthermore, population-level analysis has consistently demonstrated an increased burden of CHD in carriers of CNVs at specific genomic hotspots compared to controls, displaying the pathogenic potential of rare or *de novo* CNVs [12-14].

Bicuspid Aortic Valve (BAV) is the most common congenital heart malformation with a population prevalence of 0.5 – 2% [15]. BAV predisposes to aortic valve stenosis and thoracic aortic aneurysms and is associated with other left ventricular outflow tract lesions such as mitral valve disease and coarctation [16]. The high heritability of BAV was demonstrated in first- and second-degree relatives, who are more than ten times more likely to be diagnosed with BAV compared to matched controls [17]. BAV can occur as an isolated congenital lesion or as part of a clinical syndrome. For example, the prevalence of BAV is increased in Velocardiofacial, Loeys-Dietz, Kabuki, and Turner syndromes. Pathogenic variants of several genes are implicated in familial non-syndromic BAV, which is typically inherited as an autosomal dominant trait with reduced penetrance and variable expressivity. There is strong cumulative evidence that *GATA4, GATA6, NOTCH1, ROBO4, SMAD4*, *MUC4*, and *SMAD6* each contribute to a small percentage of non-syndromic BAV cases. Phenotypic expression of BAV disease ranges from incidental discovery in late adulthood to neonatal or childhood onset of complications. In comparison to patients with later disease onset, younger BAV cohorts tend to present with syndromic features or complex congenital malformations that are more likely to have a genetic cause, thereby increasing the power of association studies to discover clinically relevant CNVs [18]. Recently, we identified recurrent rare CNVs that were enriched for cardiac developmental genes in a young cohort with early-onset thoracic aortic aneurysms or acute aortic dissections [19].

We hypothesize that large rare genomic CNVs contribute to early onset complications of BAV. Consistent with previous observations, we predict that the burden and penetrance of rare CNVs will be increased in individuals with early onset disease when compared to elderly sporadic BAV cases and population controls. Identification of novel pathogenic CNVs can provide new insights into the genetic complexity of BAV and may be useful for personalized risk stratification or clinical guidance based on the specific recurrent CNV [20]. Therefore, we set out to describe the burden and penetrance of rare CNVs in a young cohort with early onset complications of BAV disease (EBAV).

## Materials and Methods

The study protocol was approved by the Committee for the Protection of Human Subjects at the University of Texas Health Science Center at Houston (HSC-MS-11-0185). After written informed consent, we enrolled 272 probands of European ancestry with early onset BAV disease (EBAV), which we defined as individuals with BAV who were under the age of 30 at the time of first clinical event. Clinical events were defined as aortic replacement, aortic valve surgery, aortic dissection, moderate or severe aortic stenosis or aortic regurgitation, large aneurysm (Z > 4.5), or intervention for BAV-related conditions. Those with hypoplastic left heart, known genetic mutations, genetic syndromes, or complex congenital heart disease were excluded. Affected and unaffected family members of probands were included in this cohort for a total of 544 individuals in 293 families (26 trios and 16 multiplex families). Samples were collected and genotyped similar to our previous study [21]. For comparison, we analyzed a cohort of older individuals of European ancestry with sporadic BAV disease selected from the International BAV Consortium (Table 1) [22].

**Table 1.** Summary of Case Cohorts.

| Cohort | Source | Sample Size | Array |
| --- | --- | --- | --- |
| **EBAV** | UTHealth Houston | 544 | Illumina GSA-24v1.0/2.0 |
| **BAVGWAS** | International BAV Consortium | 5040 | Illumina GSA-24v3.0 |

Cohort: name of case cohort; EBAV: family-based cohort selected for early onset complications of bicuspid aortic valve (BAV); BAVGWAS: unrelated probands with sporadic BAV disease. Source: origin of genotypes; Array: microarray used for genotyping.

Phenotypes were derived from record review with confirmation of image data whenever possible [23-24]. The computational pipeline for CNV analysis of Illumina single nucleotide polymorphism (SNP) array data included three independent CNV detection algorithms (Fig 1).

**Fig 1.**
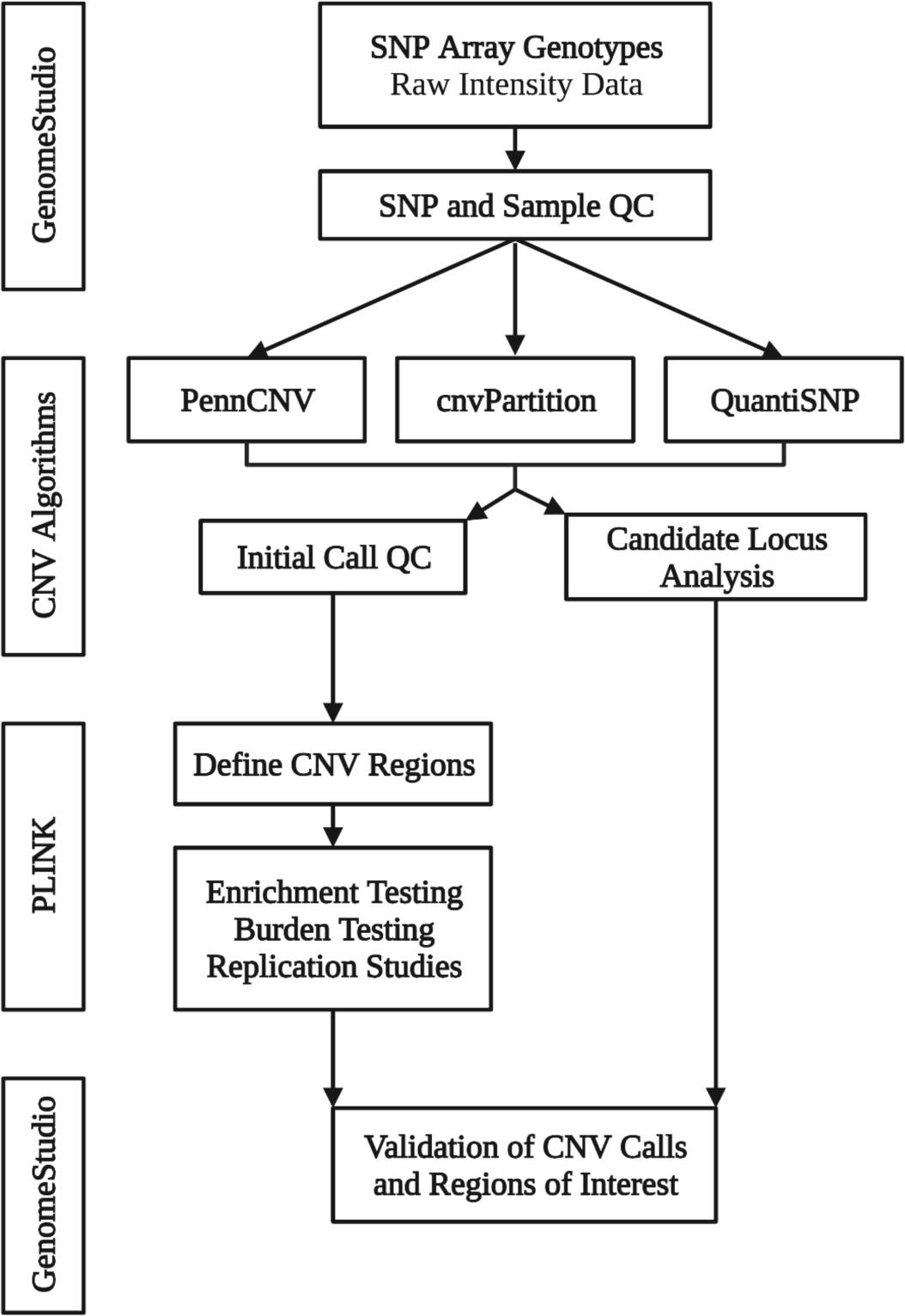
Overview of Pipeline for CNV Identification and Validation. SNP, single nucleotide polymorphism. QC, Quality control. CNV, copy number variant. The software and algorithms used for the analysis are provided in boxes to the left of the corresponding steps. Illumina raw signal intensity data was trimmed and exported using GenomeStudio. The intensity data was then analyzed with three different CNV calling algorithms (PennCNV [25], cnvPartition, and QuantiSNP [26]) to generate initial CNV calls and sample-level statistics. Sample-level quality control analysis was performed using PennCNV. PLINK [27] toolset was used to define CNV regions from initial CNV calls for subsequent burden testing, enrichment studies, and replication studies. The initial CNV calls were individually screened for CNVs intersecting with candidate loci, which we defined as genes implicated in bicuspid aortic valve disease and those discovered in our enrichment studies. CNVs of interest were then validated in GenomeStudio.

GenomeStudio was used to exclude samples with indeterminate sex or more than 5% missing genotypes, and single nucleotide polymorphisms (SNPs) with GenTrain = 0. Principal component analysis was used to remove outliers that did not cluster with European ancestry. Only SNPs common to all microarray platforms were included.

Three independent algorithms (PennCNV, cnvPartition, and QuantiSNP) were used to generate CNV calls and sample-level quality statistics from SNP intensity data. PennCNV and QuantiSNP were run on Unix clusters and cnvPartition data were exported from GenomeStudio. The analysis was run using default configurations.

PennCNV was used to generate QC data and remove CNV calls that intersect with polymorphic genomic regions. Samples that met any of the following criteria were excluded: standard deviation of the LogR ratio (obtained from PennCNV) > 0.35 or number of CNVs > 2 standard deviations above the mean for each data set. CNV calls less than 20 kilobase pairs and/or spanned by less than 6 SNP probes were excluded. The overlap function for rare CNVs in PLINK was used to construct CNV regions (CNVRs) and adjacent regions were merged using PennCNV.

LogR ratio (LRR) and B allele frequency (BAF) data at CNVRs and calls of interest were visualized in GenomeStudio for validation. For segregation analysis, GenomeStudio was used to determine the presence of CNVs in relatives.

A total of 22,014 unselected control Illumina Genotypes obtained from the Database of Genotypes and Phenotypes were analyzed using identical methods (Table in S1Table). Cohorts were paired as follows for case-control analysis based on the concordance of sample-level quality control statistics (mean number of CNV calls and mean standard deviation of the LogR Ratio): EBAV and WLS, BAVGWAS and HRS.

PLINK was used to catalog CNV calls and perform burden and enrichment studies. Case - control burden tests were restricted to large (250 - 5000 kilobase pairs), rare (occurring in less than 1 in 1000 samples; total of cases and controls), and validated CNV calls in EBAV probands. Genome Reference Consortium Human Build 37 [28] was used for CNV annotation.

## Results

Compared to BAVGWAS probands, EBAV probands were significantly younger at diagnosis, had more frequent co-existing congenital heart and vascular lesions, and underwent more frequent valve or aortic operations. A phenotype summary of the EBAV and BAVGWAS Cohorts is provided in Table 2.

**Table 2.** Characteristics of EBAV and BAVGWAS Probands.

|  | EBAV (n = 279) | BAVGWAS (n = 3141) |
| --- | --- | --- |
| Female (%) | 33 | 29 |
| Age at diagnosis (years) | 17 □ 13 | 52 □ 16 |
| TAA (%) | 20 | 37 |
| Predominant AR (%) | 12 | 40 |
| Predominant AS (%) | 20 | 37 |
| Other Lesions (%) | 53 | 1 |
| Aortic Replacement (%) | 27 | 16 |
| Aortic Valve Surgery (%) | 40 | 16 |

N: number of cases; □ , standard deviation; TAA, thoracic aortic aneurysm; AR: aortic regurgitation; AS, aortic stenosis; Other Lesions, other congenital heart malformations (primarily coarctation or ventricular septal defect). We had phenotype information for 279 EBAV probands but did not have access to genotype information for all samples.

CNV analysis is summarized in Table 3. The percentages of individuals with large and rare CNV regions were relatively consistent throughout datasets. The prevalence of large and rare CNVs, specifically large genomic deletions, was increased in EBAV cases compared to controls (Table S2).

**Table 3.**
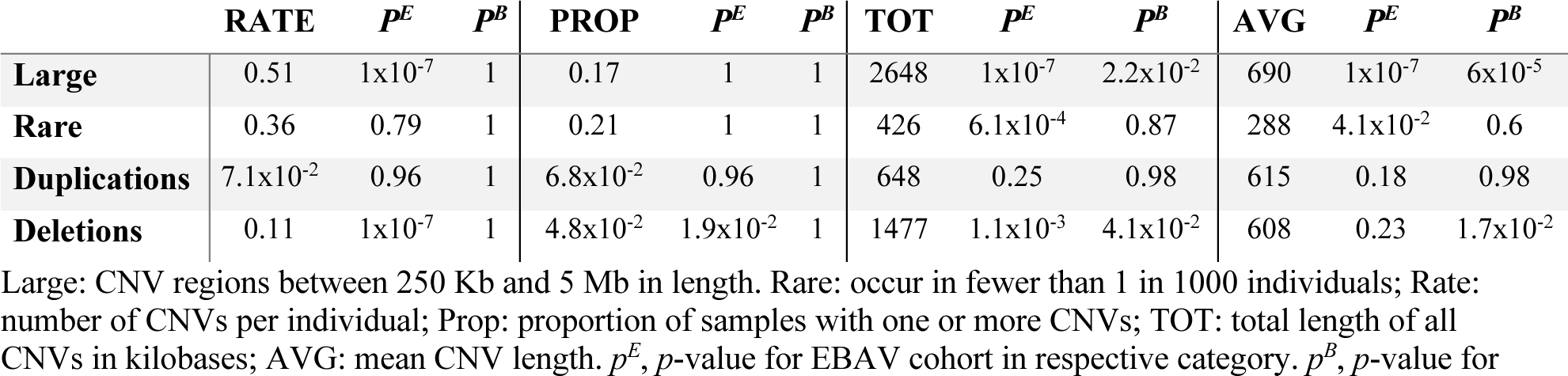
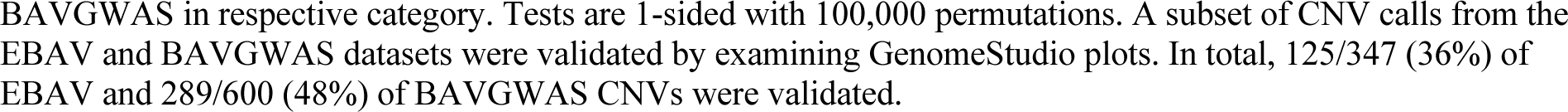
Summary of CNV Calls for EBAV Cohort.

| | RATE | $p^E$ | $p^B$ | PROP | $p^E$ | $p^B$ | TOT | $p^E$ | $p^B$ | AVG | $p^E$ | $p^B$ |
| --- | --- | --- | --- | --- | --- | --- | --- | --- | --- | --- | --- | --- |
| Large | 0.51 | $1 \times 10^{-7}$ | 1 | 0.17 | 1 | 1 | 2648 | $1 \times 10^{-7}$ | $2.2 \times 10^{-2}$ | 690 | $1 \times 10^{-7}$ | $6 \times 10^{-5}$ |
| Rare | 0.36 | 0.79 | 1 | 0.21 | 1 | 1 | 426 | $6.1 \times 10^{-4}$ | 0.87 | 288 | $4.1 \times 10^{-2}$ | 0.6 |
| Duplications | $7.1 \times 10^{-2}$ | 0.96 | 1 | $6.8 \times 10^{-2}$ | 0.96 | 1 | 648 | 0.25 | 0.98 | 615 | 0.18 | 0.98 |
| Deletions | 0.11 | $1 \times 10^{-7}$ | 1 | $4.8 \times 10^{-2}$ | $1.9 \times 10^{-2}$ | 1 | 1477 | $1.1 \times 10^{-3}$ | $4.1 \times 10^{-2}$ | 608 | 0.23 | $1.7 \times 10^{-2}$ |

Large: CNV regions between 250 Kb and 5 Mb in length. Rare: occur in fewer than 1 in 1000 individuals; Rate: number of CNVs per individual; Prop: proportion of samples with one or more CNVs; TOT: total length of all CNVs in kilobases; AVG: mean CNV length. $p^E$ , $p$ -value for EBAV cohort in respective category. $p^B$ , $p$ -value for
BAVGWAS in respective category. Tests are 1-sided with 100,000 permutations. A subset of CNV calls from the EBAV and BAVGWAS datasets were validated by examining GenomeStudio plots. In total, 125/347 (36%) of EBAV and 289/600 (48%) of BAVGWAS CNVs were validated.

There were 34 large (>250 Kb), rare (<1:1000 in dbGAP controls) CNV regions that involved protein-coding genes in EBAV cases (Table S3). Seven of these genic CNVs were enriched in EBAV cases compared to WLS controls with a genome-wide adjusted empiric P < 0.05. These CNVs included the genes *PCP4*, *DSCAM*, *MIR4760*, and *DSCAM-AS1* in 21q22 and *GATA4*, *C8orf49*, *NEIL2*, *FDFT1*, and *CTSB* in 8p23. Large duplications involving the Velocardiofacial (VCFS) region in 22q11.2 and 1q21.1 microduplications were also enriched in EBAV cases (Table S4). The overall burden of large, rare, genic CNVs was not different between EBAV cases and WLS controls. However, the burden of large, rare genic CNVs intersecting with genes known to cause BAV when mutated or implicated in syndromic BAV was significantly increased in EBAV cases (Table 4).

**Table 4.** Burden Testing of Rare EBAV CNVs.

|  | <u>EBAV</u> |  | <u>WLS</u> |  | RR | *P* |
| --- | --- | --- | --- | --- | --- | --- |
|  | Calls | Rate | Calls | Rate |  |  |
| **Genic** | 28 | 0.8 | 1151 | 0.65 | 1.2 | 0.23 |
| **Deletions** | 11 | $3.8 \times 10^{-2}$ | 439 | $4.6 \times 10^{-2}$ | 0.81 | 0.78 |
| **BAV** | 3 | $1.0 \times 10^{-2}$ | 1 | $1.1 \times 10^{-4}$ | 97 | $1.1 \times 10^{-3}$ |
| **Total** | 34 | - | 1443 | - | - | - |

Calls: total number of CNVs that met the specified criteria. Rate: number of CNVs per individual; RR: relative risk; *P*: p-value; Genic: CNVs that intersect with genes; BAV: CNVs that intersect with genes that are known to cause bicuspid aortic valve (BAV) when mutated or implicated in syndromic BAV. Total: total number of large, rare CNVs or CNVRs. Tests are 2-sided using 100,000 permutations.

We also scrutinized genomic regions that are implicated in CHD by careful analysis of data from individual CNV algorithms to detect subtle copy number alterations. We identified additional rare EBAV CNVs that intersect with CHD candidate genes *CELSR1*, *GJA5*, *RAF1*, *LTBP1, KIF1A*, *MYH11*, *MAPK3*, *TTN*, and the VCFS region in 22q11.2. We detected additional *GATA4* and *DSCAM* CNVs in multiplex families. These CNVs were enriched in EBAV cases compared to WLS controls (Table 5).

**Table 5.** CNVs Affecting Congenital Heart Disease Genes in EBAV Cohort.

| Region | Genes | Case | Control | OR | 95% CI |
| --- | --- | --- | --- | --- | --- |
| Chr22:46261909-51187440 | *CELSR1* | 1 | 1 | 33 | 2.1 to 530 |
| Chr1:146326373-147340734 | *GJA5* | 1 | 2 | 17 | 1.5 to 183 |
| Chr3:12599717-12803792 | *RAF1* | 1 | 2 | 17 | 1.5 to 183 |
| Chr22:41278694-41813285 | *DSCAM* | 4 | 2 | 67 | 12 to 367 |
| Chr8:11495032-11856903 | *GATA4* | 4 | 0 | 301 | 16 to 5599 |
| Chr22:19000000-22000000 | *TBX1, CRKL* | 4 | 10 | 13 | 4.2 to 43 |
| Chr16:15484868-16295863 | *MYH11* | 2 | 22 | 3.0 | 0.70 to 13 |
| Chr2:241652252-241678528 | *KIF1A* | 3 | 22 | 4.5 | 1.3 to 15 |
| Chr2:32775984-33331219 | *LTBP1* | 2 | 26 | 2.5 | 0.60 to 11 |

Region: coordinates corresponding to the minimum overlap region of CNVs; Genes: cardio-developmental candidate genes in the region. Case: number of large and rare CNVs in EBAV cases that intersect with region of interest. Control: number of CNVs in WLS cohort that intersect with region of interest. OR: odds ratio; 95% CI, 95% confidence interval for respective odds ratio.

Next, we attempted to replicate our observations by identifying CNVs in the BAVGWAS dataset that overlapped with rare EBAV CNVs. We found that large duplications involving *SOX7* and *GATA4* in 8p23 and the VCFS region in 22q11.2 were also significantly enriched in BAVGWAS cases compared to HRS controls (Table 6, Table S6 and S7).

**Table 6.** CNVs Affecting Congenital Heart Disease Genes in BAVGWAS Cohort.

| Region | Genes | Case | Control | OR | 95% CI |
| --- | --- | --- | --- | --- | --- |
| Chr3:29993977-31273870 | *TGFBR2* | 1 | 0 | 5.6 | 0.23 to 138 |
| Chr9:101861767-102092282 | *TGFBR1* | 1 | 0 | 5.6 | 0.23 to 138 |
| Chr21:41577819-41842252 | *DSCAM* | 2 | 1 | 3.7 | 0.34 to 41 |
| Chr22:46924254-46931077 | *CELSR1* | 3 | 1 | 5.6 | 0.58 to 54 |
| Chr2:111404636-11310378 | *TMEM87B, FBLN7* | 3 | 2 | 2.8 | 0.47 to 17 |
| Chr8:11385469-11821835 | *GATA4* | 8 | 1 | 15 | 1.9 to 120 |
| Chr12:7918339-8130958 | *NANOG* | 10 | 2 | 9.4 | 2.1 to 43 |
| Chr2:147166377-147308112 | *GJA5* | 4 | 10 | 0.75 | 0.23 to 2.4 |
| Chr16:29664753-30199713 | *MAPK3* | 3 | 15 | 0.37 | 0.11 to 1.3 |
| Chr22:19000000-22000000 | *TBX1, CRKL* | 18 | 11 | 3.1 | 1.4 to 6.5 |
| Chr2:32689829-33299434 | *LTBP1* | 9 | 22 | 0.76 | 0.35 to 1.7 |
| Chr16:15240816-16281154 | *MYH11* | 13 | 27 | 0.90 | 0.46 to 1.7 |
| Chr2:241640262-241689833 | *KIF1A* | 13 | 30 | 0.81 | 0.42 to 1.6 |

Region: coordinates corresponding to the minimum overlap region of CNVs; Genes: cardio-developmental candidate genes in the region. Case: number of large and rare CNVs in BAVGWAS cases that intersect with region of interest. Control: number of CNVs in HRS cohort that intersect with region of interest. OR: odds ratio; 95% CI, 95% confidence interval for respective odds ratio.

CNVs intersecting with *GATA4* and *DSCAM* significantly overlapped between EBAV and BAVGWAS datasets (Fig 2). On average, the *GATA4* CNVs were larger in the BAVGWAS dataset while the *DSCAM* CNVs were larger in the EBAV dataset.

**Fig 2.**
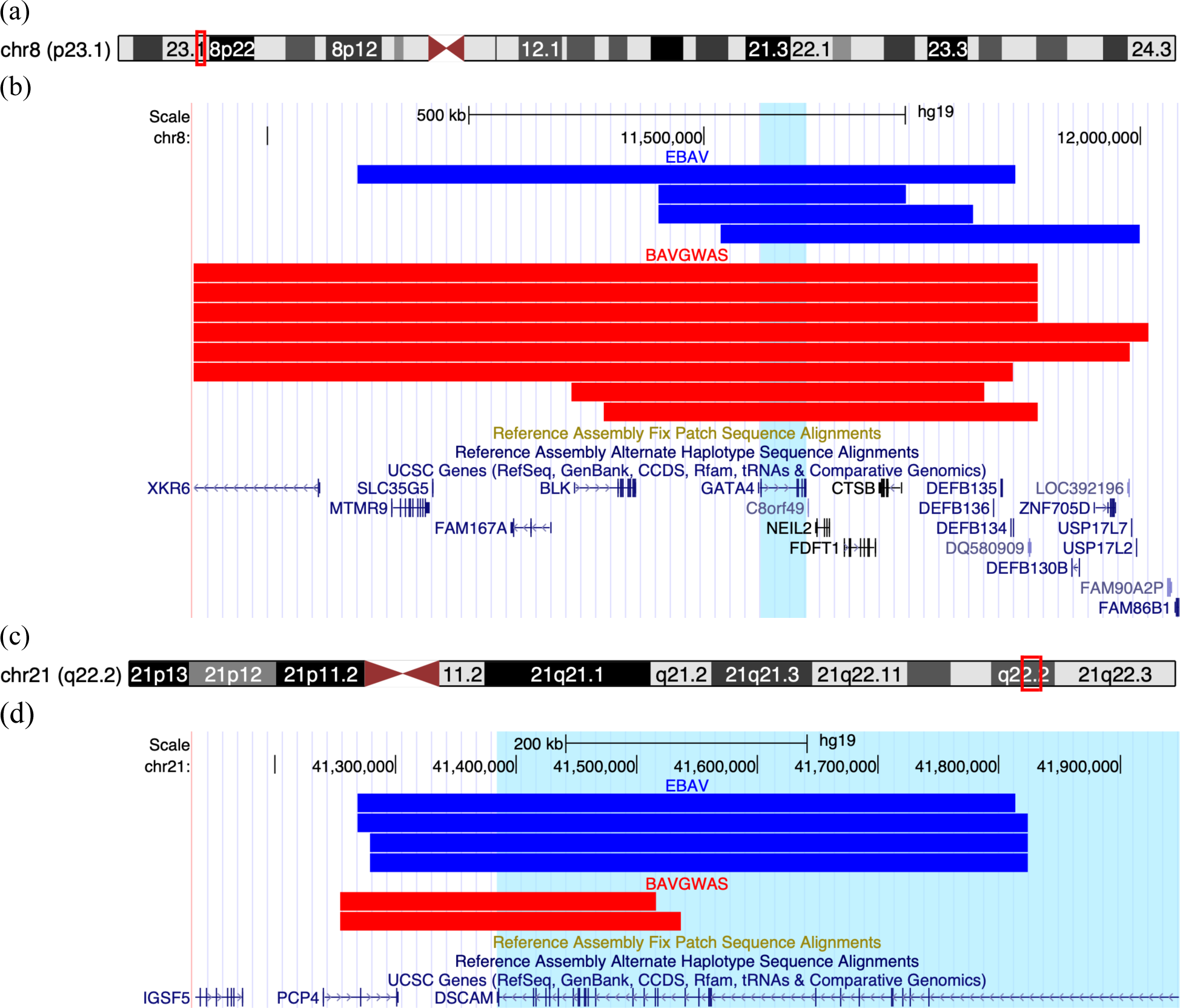
UCSC Genome Browser Plots of *GATA4* and *DSCAM* Variants. (a) Ideogram of Chromosome 8 with view of image in (b) outlined in red box. (b) Plot of *GATA4* variants. Each bar represents a copy number variant (CNV). CNVs from the EBAV cohort are in blue and CNVs from the BAVGWAS cohort are in red. The region spanned by *GATA4* has been highlighted in light blue. (c) Ideogram of Chromosome 21 with view of image in (d) outlined in red box. (d) Plot of *DSCAM* variants. EBAV CNVs are in blue and BAVGWAS CNVs are in red. The region spanned by *DSCAM* is highlighted in light blue. Figures constructed using the UCSC Genome Browser, http://genome.ucsc.edu [29].

We identified 7 additional CNV regions that are enriched in BAVGWAS cases but not in EBAV and are rare or absent in controls (Table S5). *NANOG* and *NIBPL* are essential for early heart development, and mutation of *NIBPL* causes Cornelia-de Lange syndrome with a spectrum of congenital heart malformations including BAV.

We also identified 21 very large genomic CNVs more than 5 Mb in length in the BAVGWAS dataset. Analysis of GenomeStudio data showed that most of these were mosaic loss of heterozygosity regions or duplications. Nine were large germline chromosome-scale aberrations, including two cases of trisomy 21 (Table S8). We did not identify any large X chromosome copy variants that may be consistent with Turner syndrome. There were no megabase-scale copy number variants in the EBAV dataset.

Pedigree analysis showed that several CNVs involving *CELSR1*, *LTBP1*, *KIF1A*, *GATA4*, and *DSCAM* segregate with BAV in EBAV families (Table S9). CNV carriers tended to present due to moderate or severe aortic regurgitation requiring valvular surgery. One proband had aortic coarctation. The youngest age at presentation was 13 years. There were no sex differences in presentation between CNV carriers.

## Discussion

We identified large, rare, and likely pathogenic CNVs in almost 10% of EBAV probands that are enriched in genes that cause BAV when mutated. The percentage of EBAV cases with likely pathogenic CNVs is similar to our previous observations in a cohort with early onset TAD [30]. Enrichment of CNVs involving *GATA4* and *DSCAM* in EBAV cases replicated in two additional BAV datasets and thousands of unselected control genotypes. This analysis provides compelling evidence that rare CNVs collectively cause more BAV cases than any single mutated gene.

GATA-Binding Protein 4 is a transcription factor that is required for cardiac and neuronal differentiation during embryogenesis [31]. Mutations of *GATA4* and its homologs *GATA5* and *GATA6* cause congenital heart lesions [32]. Mutations in the *GATA4* gene have been linked to a range of congenital heart diseases in humans, such as cardiac septal defects, tetralogy of Fallot, amd patent ductus arteriosus [33]. Patients with BAV who have rare functional variants in the *GATA* family exhibit varying degrees of aortopathy expression, including aortic aneurysm, dissection, and/or aortic stenosis. Alonso-Montes et al. described 4 predicted deleterious *GATA4* mutations in 122 non-syndromic BAV probands who did not have affected relatives [34]. Rare *GATA4* deletions and putative loss of function mutations are also implicated in CHD with distinctive features, underlining the importance of *GATA4* dosage to cardiac development [35-36]. Glessner et al. discovered large *de novo* (∼4Mb) duplications involving *GATA4* in CHD trios with conotruncal defects or left ventricular outflow tract obstructive lesions [37]. Some duplications were inherited from apparently unaffected parents. Zogopoulos and Yu described similar genomic duplications in unaffected individuals and in unselected control genotypes [38-39].

These observations are consistent with low-penetrance CHD in *GATA4* duplication carriers. Similar to other complex and multifactorial disorders, CHD pathogenesis is likely caused by the cumulative impact of multiple CNVs or mutations, each exerting small to moderate effects to collectively disrupt cardiac development. For example, the frequency of congenital heart lesions is increased in individuals with velocardiofacial syndrome who have 22q1.2 deletions and a common 12p13.31 duplication involving the *SLC2A3* gene. The *SLC2A3* CNV likely functions as a modifier of the cardiac phenotype associated with 22q11 deletion syndrome, exemplifying a “two-hit” model [40].

More than half of patients with Down syndrome have congenital heart malformations due to the interaction of multiple dosage-sensitive CHD genes on chromosome 21 [41-43]. Down syndrome cell adhesion molecule, previously shown to play a critical role in neurogenesis, has also been implicated in the pathophysiology of CHD [44]. Analysis of rare segmental trisomies of chromosome 21 suggested that duplication of *DSCAM* and the contiguous *COL6A1* and *COL6A2* genes may cause septal abnormalities and other Down Syndrome-related CHD lesions, including BAV. Overexpression of *DSCAM* and *COL6A2* causes cardiac malformations in mice [45]. Our findings suggest that rare CNVs involving *DSCAM* may contribute to some non-syndromic BAV cases.

Consistent with previous observations, *GATA4* and *DSCAM* CNVs segregated with disease in multiple families, but are not fully penetrant and were detected in some unaffected relatives. Intriguingly, large 22q11.2, *GATA4* and *DSCAM* CNVs were more highly enriched in EBAV than in BAVGWAS cases, suggesting that these CNVs may drive early onset BAV disease. These results are consistent with our observation that pathogenic CNVs involving candidate BAV genes are also enriched in EBAV compared to BAVGWAS cases. Our data suggests that pathogenic CNVs at these loci may predict accelerated disease onset or more severe complications.

We also identified recurrent rare CNVs of specific dosage-sensitive regions that affect cardiac developmental genes and are implicated in non-syndromic CHD. Recurrent 1q21.1 distal deletions encompassing *GJA5*, the gene encoding Connexin-40, are associated with CHD lesions including BAV. A study of 807 TOF cases showed significant enrichment of small duplications spanning the *GJA5* gene, providing compelling evidence that it acted as the primary candidate gene, supporting the association of *GJA5* and CHD [31]. Additionally, cardiac abnormalities have been documented in mice with a targeted *GJA5* deletion, implying that haploinsufficiency of *GJA5* might contribute to cardiac defects in individuals affected by 1q21.1 deletions [46]. *CELSR1*, a cadherin superfamily member, is mutated in families with BAV and hypoplastic left heart syndrome [47]. *LTBP1* encodes an extracellular matrix protein that regulates TGF-beta and fibrillin and has been implicated in congenital heart lesions [48]. *KIF1A*, encoding a kinesin microtubule transporter, was implicated in a dominant multisystem syndromic disorder with valvular and cardiac defects [49]. Mutation of *MYH11* causes familial thoracic aortic aneurysms and dissections with an increased prevalence of BAV [50]. *TTN* mutations cause dilated cardiomyopathy and are associated with other left-sided congenital lesions [51]. Mutations or copy number changes involving these genes all cause a wide spectrum of penetrance and phenotypic severity, consistent with sensitivity to genetic or clinical modifiers.

Our combinatorial analysis method eliminated many CNVs that were detected by single algorithms or did not meet quality control benchmarks. Therefore, our analysis likely underestimated the contribution of rare pathogenic CNVs to BAV. We also recognize that cardiac development involves the complex interaction of many genes. We selectively validated individual CNVs at loci of interest but may have underrepresented CNVs that had no *a priori* relationship with CHD. The apparent penetrance of some CNVs may be less than expected due to missing phenotypic information. The available clinical data was not sufficiently detailed to permit genotype-phenotype correlations with specific CHD clinical features.

In conclusion, we identified large rare CNVs in a significant proportion of BAV cases, including a subset of CNVs that may predict early onset complications of BAV disease. These observations add to the evidence that rare CNVs may eventually have clinical utility for risk stratification and personalized disease management.

## Data Availability

All data produced in the present work are contained in the manuscript.

## Acknowledgments

We thank Joana Castillo and Jacqueline Jennings for sample preparation, and Gladys Zapata, Nitesh Mehta, and the Laboratory for Translational Genomics at Baylor College of Medicine for microarray genotyping. This study was supported in part by R01HL137028 (SP). Fig 1 created with BioRender.com.

The EBAV Investigators are: Shaine A. Morris, Rita Milewski, Giuseppe Limongelli, Allesandro Della Corte, Laura Perrone, Yuli Y. Kim, Hector Michelena, Maria G. Andreassi, Arturo Evangelista, Denver Sallee, Anji Yetman, Kim McBride, Eduardo Bossone, Rodolfo Citro, Dawn S. Hui, Malenka M. Bissell, Andrea Ballotti, Ilenia Foffa, Margot De Marco, Anthony Caffarelli, Rita Weise, Julie DeBacker, Laura Muino Mosquera, Robbin Cohen, Laura Dos Subira, Justin T. Tretter, Anna Sabe Rotes, Martina Caiazza, Lamia Ait Ali, Francesca Pluchinotta, and Simon C. Body.

The BAVCon Investigators are: Simon Body, Alessandro Della Corte, Alessandro Frigiola, Andrea Ballotta, Arturo Evangelista, Betti Giusti, Bo Yang, Carlo de Vincentiis, Dan Gilon, David Messika Zeitoun, Dianna M. Milewicz, Eduardo Bossone, Eric Eisselbacher, Francesca R. Pluchinotta, Giuseppe Limongelli, Gordon S. Huggins, Hector I. Michelena, J. Daniel Muehlschelgel, Kim Eagle, Lasse Folkerson, Malenka M. Bissell, María Brion, Patrick Mathieu, Per Eriksson, Peter Lichtner, Rodolfo Citro, Ronen Durst, Sébastien Thériault, Siddharth K. Prakash, Thoralf M. Sundt, Vicenza Stefano Nistri, Yahn Bossé.

## Supplemental Data

**S1 Table.** Summary of Control Cohorts. Cohort, name of control cohort. Study, study from which genotypes were obtained. Samples, number of control samples in each dataset. Accession, Database of Genotypes and Phenotypes accession number. Microarray, Illumina microarray used for genotyping.

| Cohort | Study | Samples | Accession | Microarray |
| --- | --- | --- | --- | --- |
| **WLS** | Wisconsin Longitudinal Study on Aging | 8969 | Phs001157.v1.pl | Illumina HumanOmniExpress-24 v1.1 |
| **HRS** | Health and Retirement Study | 9426 | phs000428.v2.pl | Illumina Human Omni2.5-Quad |

**S2 Table.**
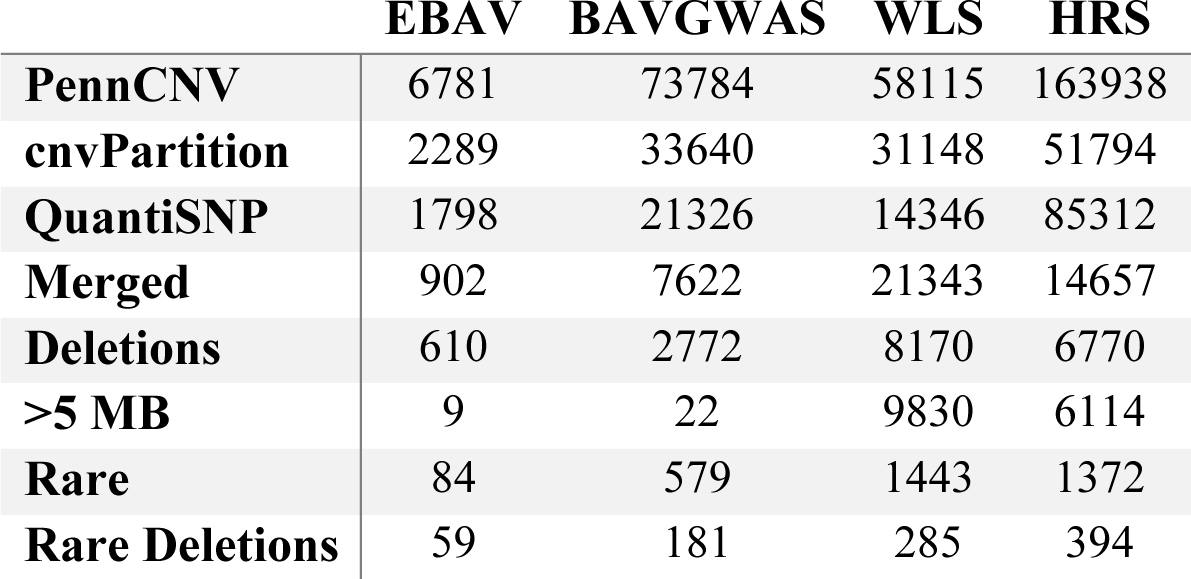
Comprehensive CNV Summary. EBAV, EBAV Cohort including cases and unaffected family members. BAVGWAS, BAVGWAS cohort. WLS, WLS cohort. HRS, HRS cohort. PennCNV, number of CNV calls detected by PennCNV algorithm after quality control. cnvPartition, number of CNV calls detected by cnvPartition algorithm after quality control. QuantiSNP, number of CNVs detected by QuantiSNP algorithm after quality control. Merged, number of CNV regions after merging initial calls. Deletions, number of CNV regions that are deletions. >5 MB, number of CNV regions that are larger than 5 megabases. Rare, number of large (> 250 kilobases and less than 5 megabases) CNV regions that occur in less than 1 in 1000 samples based on case-control cohort pairs (EBAV and WLS; BAVGWAS and HRS). R. Del., number of large, rare deletions. All values reflect the total CNV calls and regions prior to validation in GenomeStudio.

**S3 Table.**
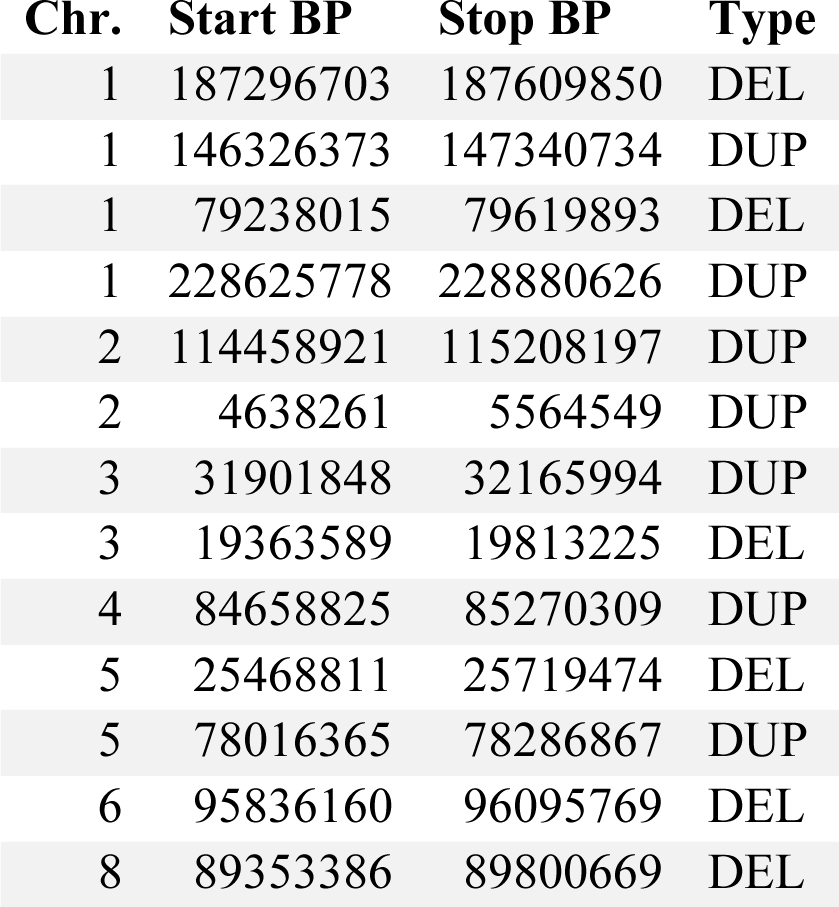
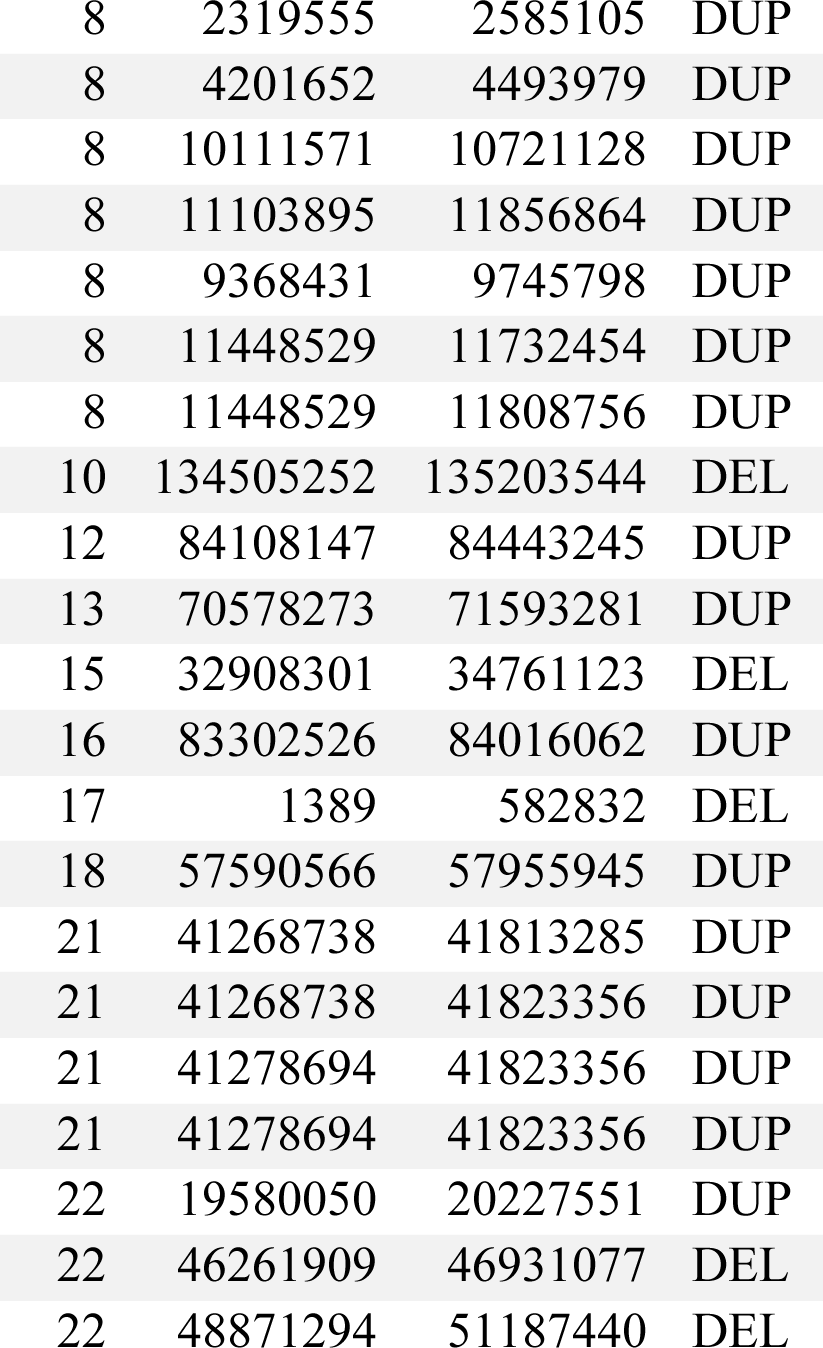
Large, Rare Copy Number Variants Identified in the EBAV Cohort. Chr., Chromosome on which CNV is located. Start BP, start basepair of CNV. Stop BP, stop basepair of CNV. Type, denotes if a CNV is a duplication (DUP) or deletion (DEL) event. All CNVs were validated in GenomeStudio.

**S4 Table.**
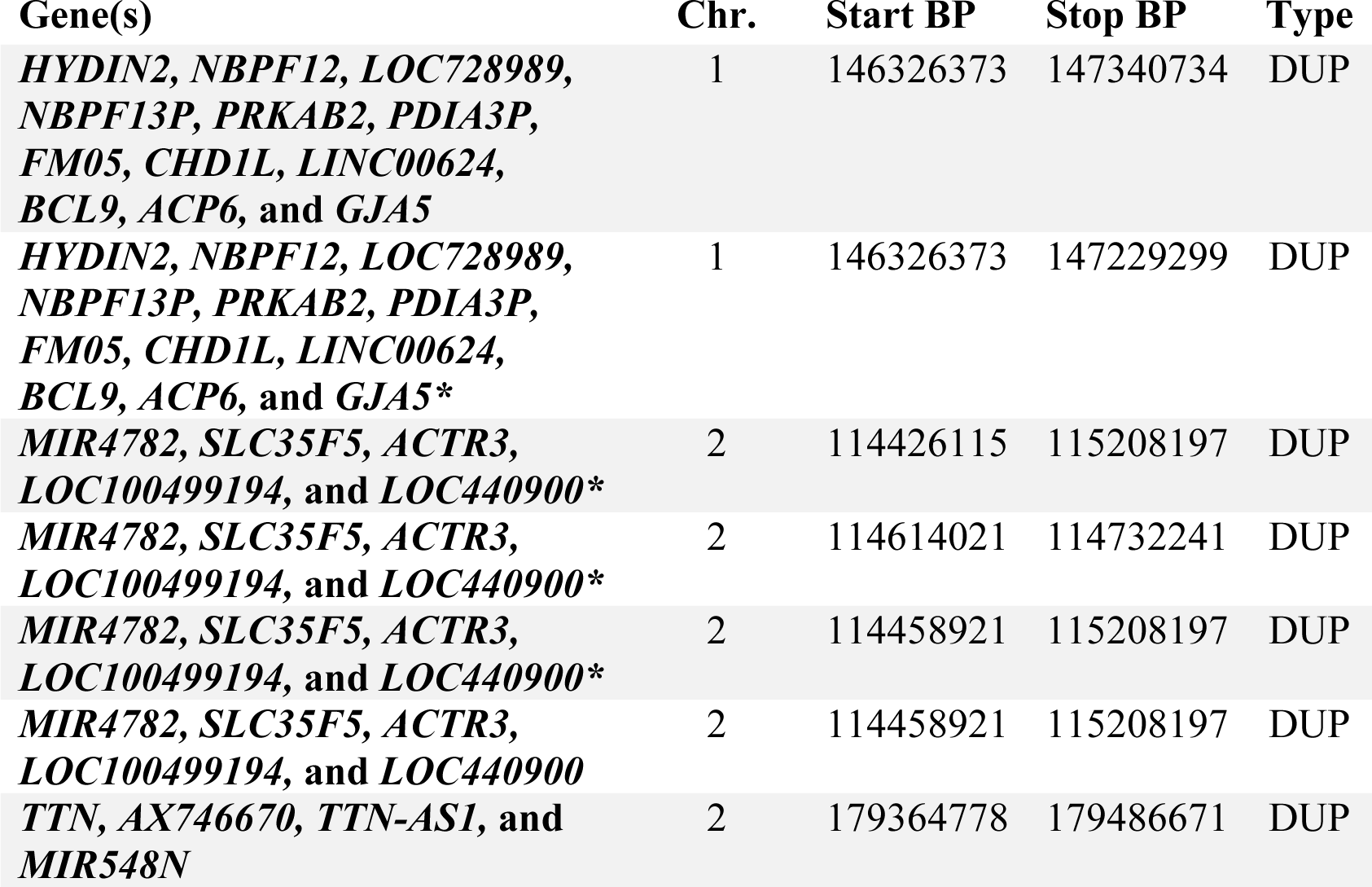
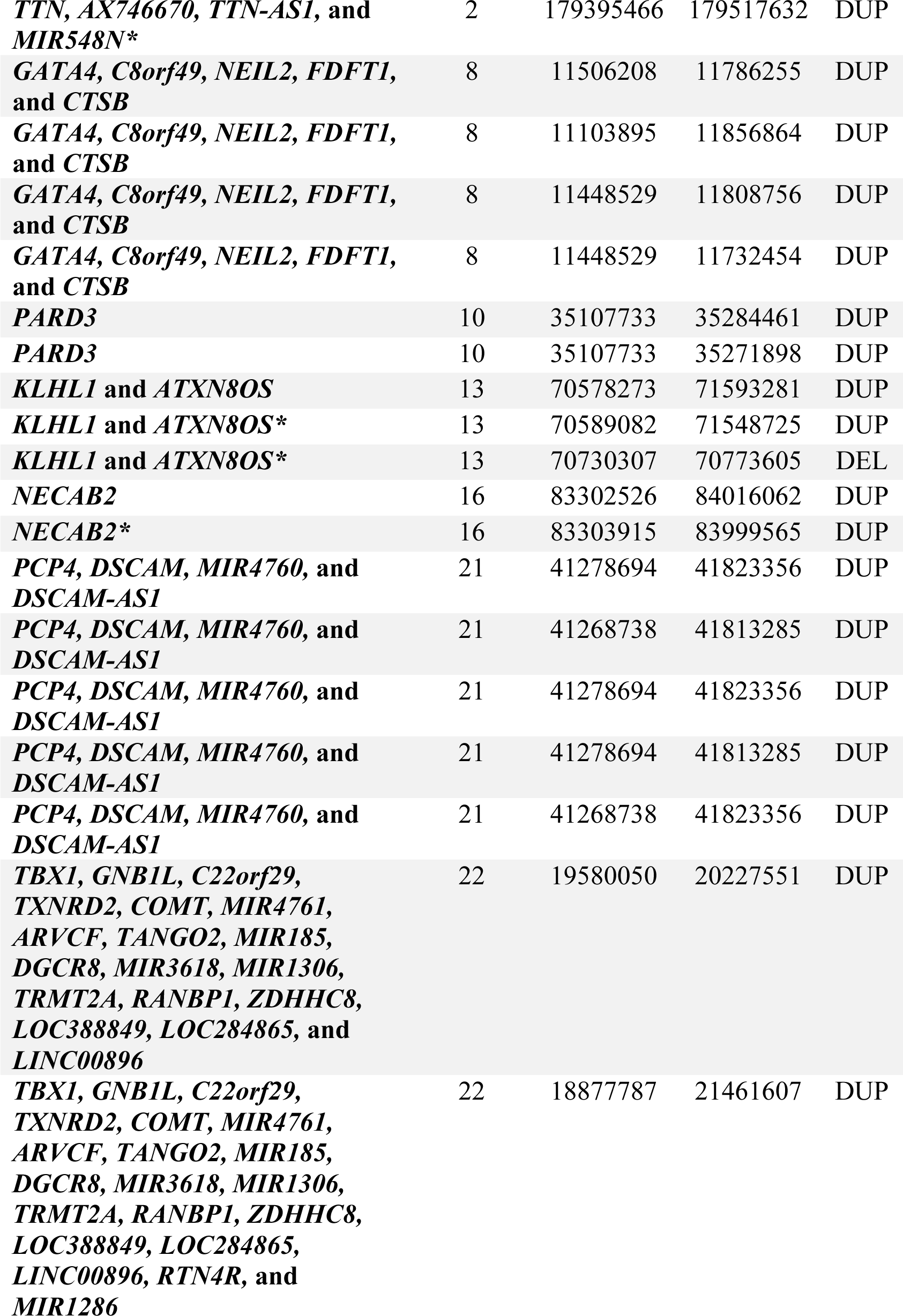
Rare CNVs Enriched in EBAV Cohort. Gene(s), genes intersected by CNV. Chr, chromosome on which each CNV is on. Start BP, start basepair of each CNV. Stop BP, stop basepair of each CNV. Type, denotes if a CNV was a duplication (DUP) or deletion (DEL) event. * Indicates the call was from an unaffected family member.

**S5 Table.**
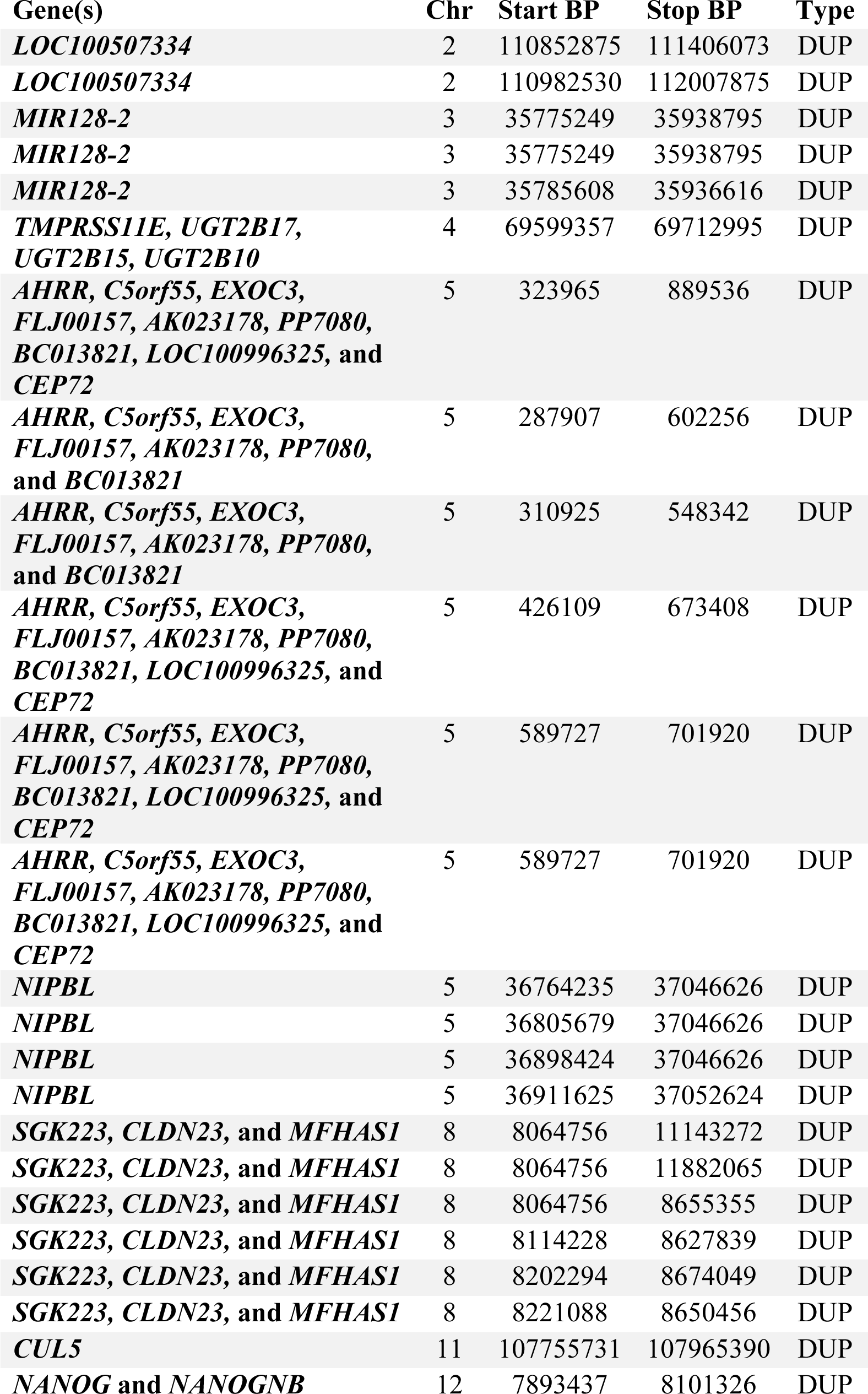
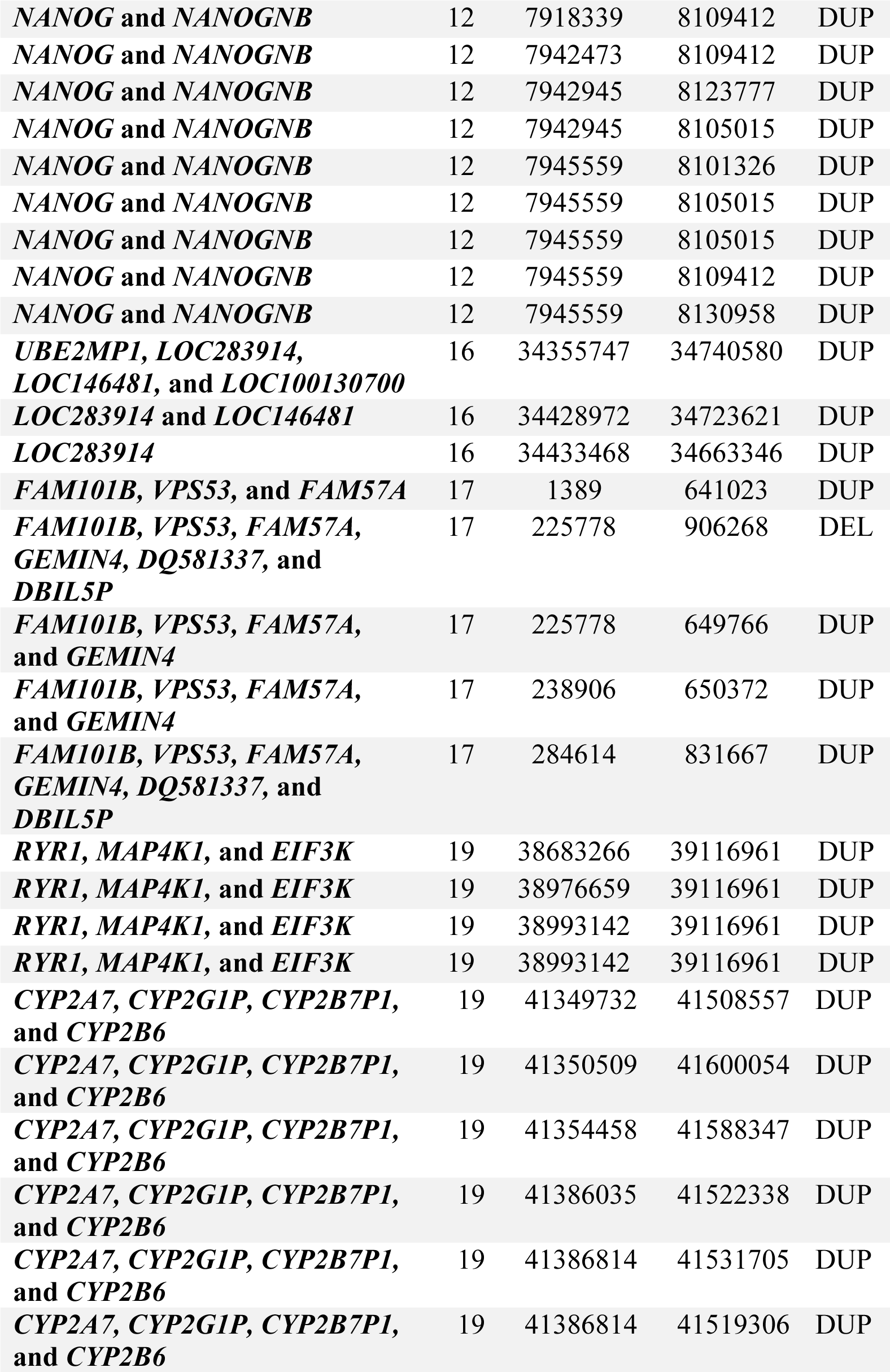
Rare CNVs Enriched in BAVGWAS Cohort. Gene(s), genes intersected by CNV. Chr, chromosome on which each CNV is on. Start BP, start basepair of each CNV. Stop BP, stop basepair of each CNV. Type, denotes if a CNV was a duplication (DUP) or deletion (DEL) event.

**S6 Table.**
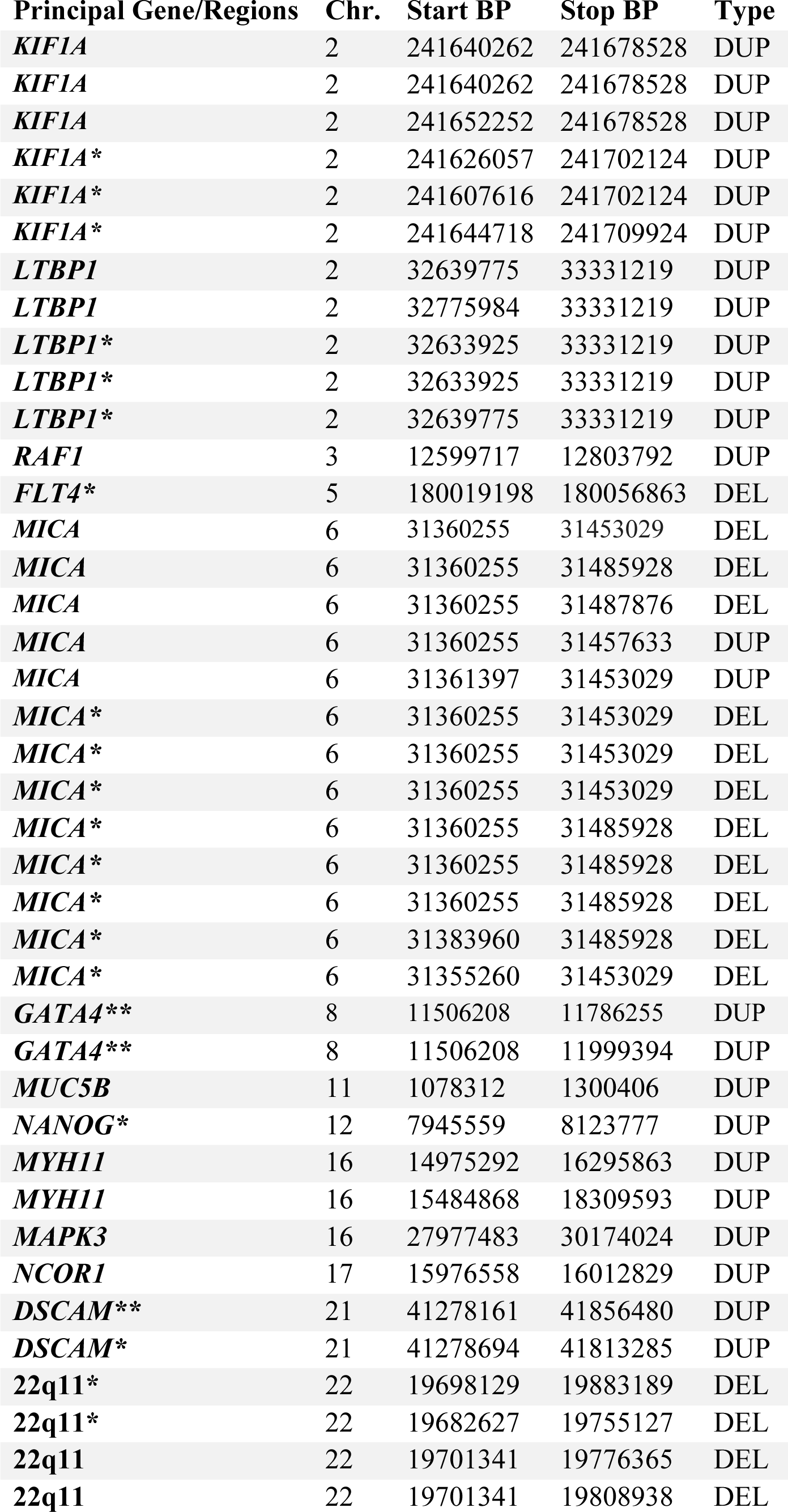
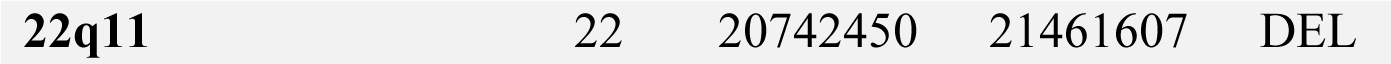
EBAV CNVs intersecting with Genes of Interest. Gene/Region, Principal gene or region of interest intersected by CNV. Chr, chromosome on which each CNV is on. Start BP, start basepair of each CNV. Stop BP, stop basepair of each CNV. Type, denotes if a CNV was a duplication (DUP) or deletion (DEL) event. * Indicates the call was from an unaffected family member. ** Indicates the call was from an affected family member from a multiplex family.

**S7 Table.**
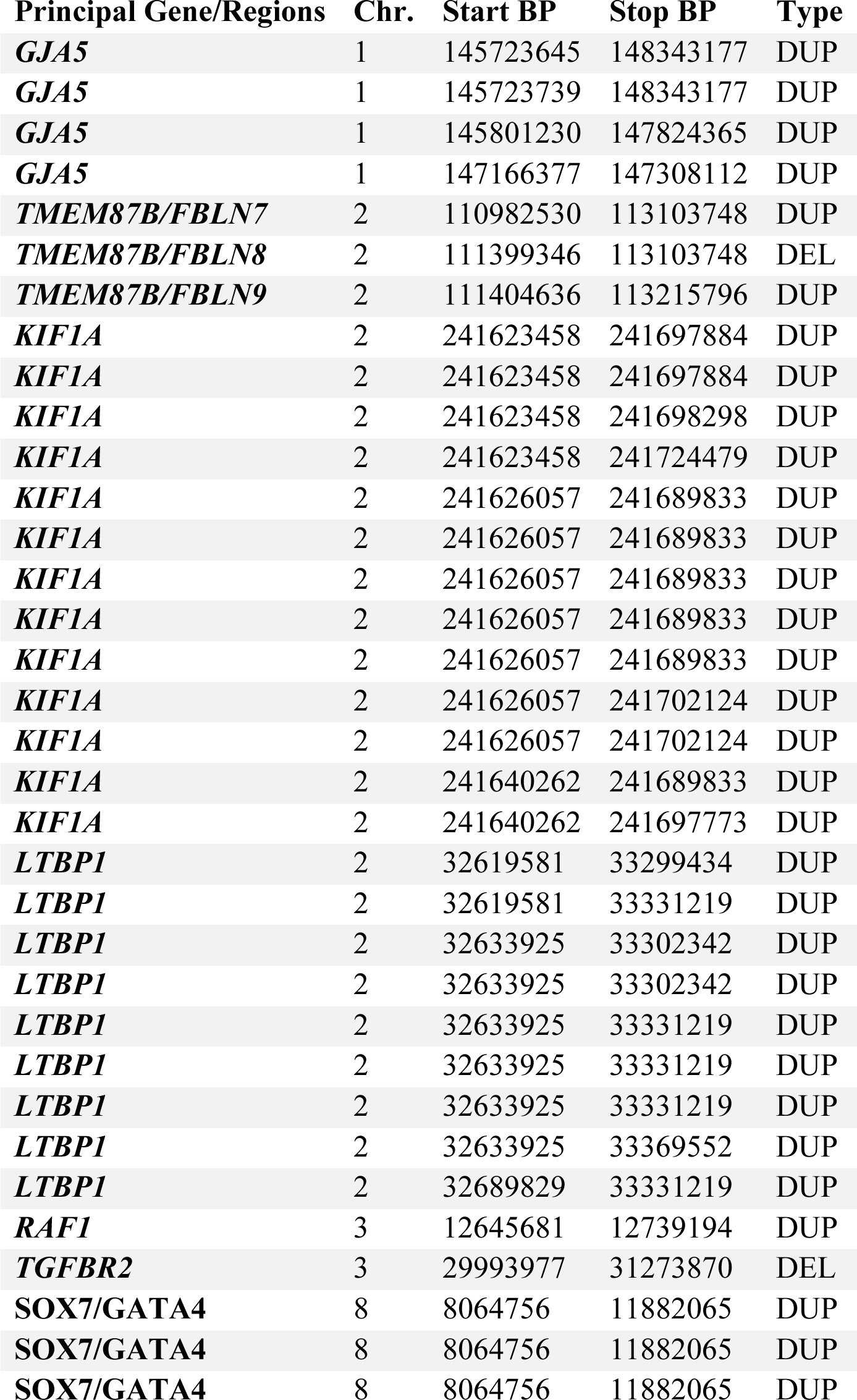
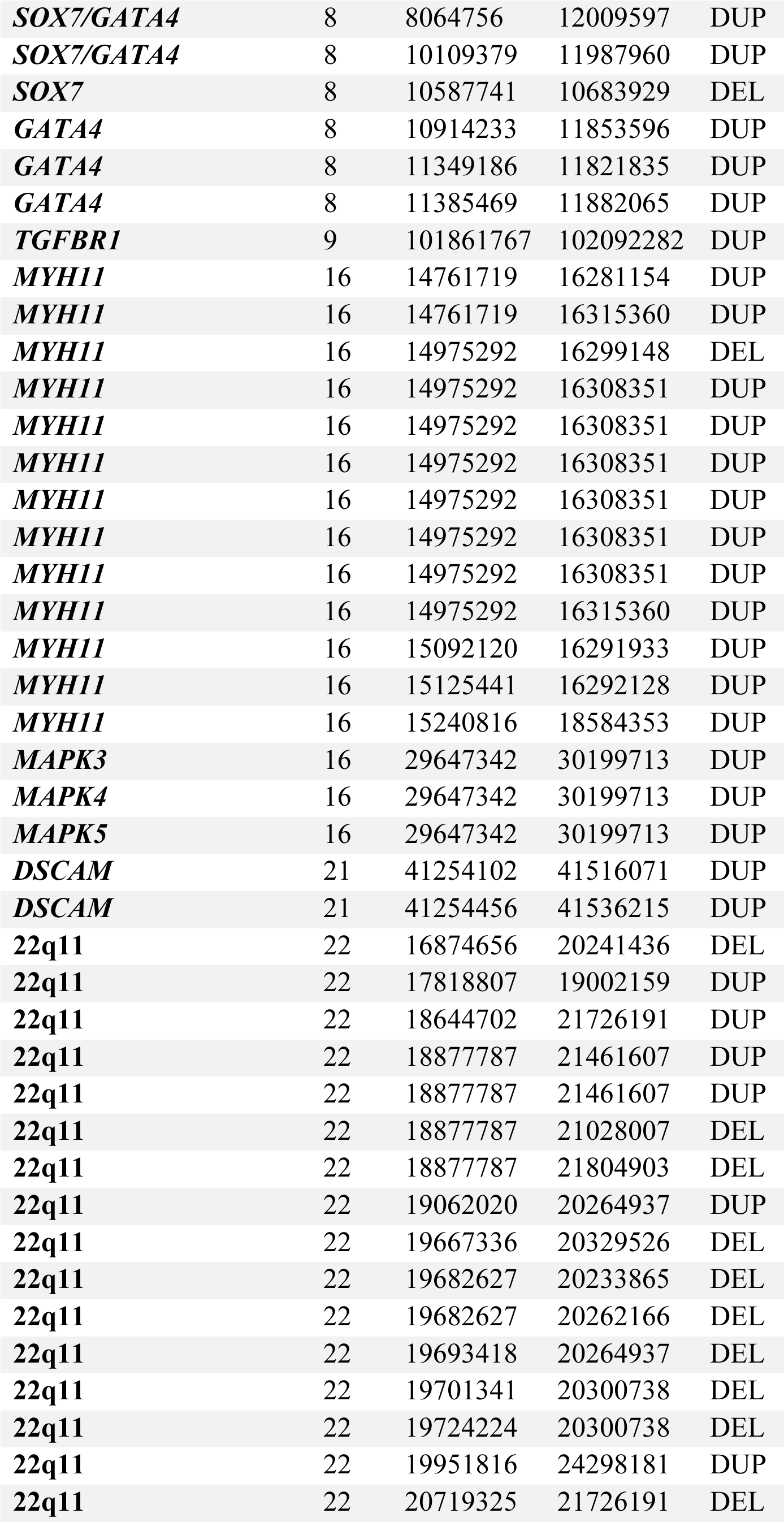
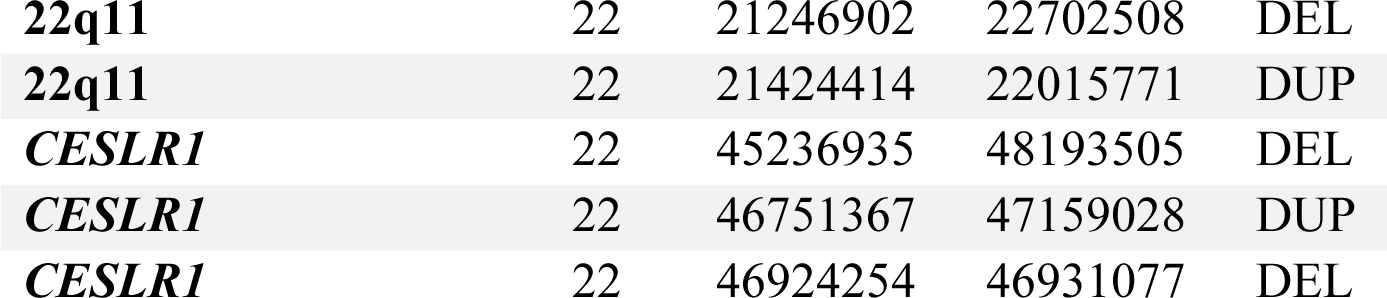
BAVGWAS CNVs intersecting with Genes of Interest. Gene/Region, Principal gene or region of interest intersected by CNV. Chr, chromosome on which each CNV is on. Start BP, start basepair of each CNV. Stop BP, stop basepair of each CNV. Type, denotes if a CNV was a duplication (DUP) or deletion (DEL) event.

**S8 Table.**
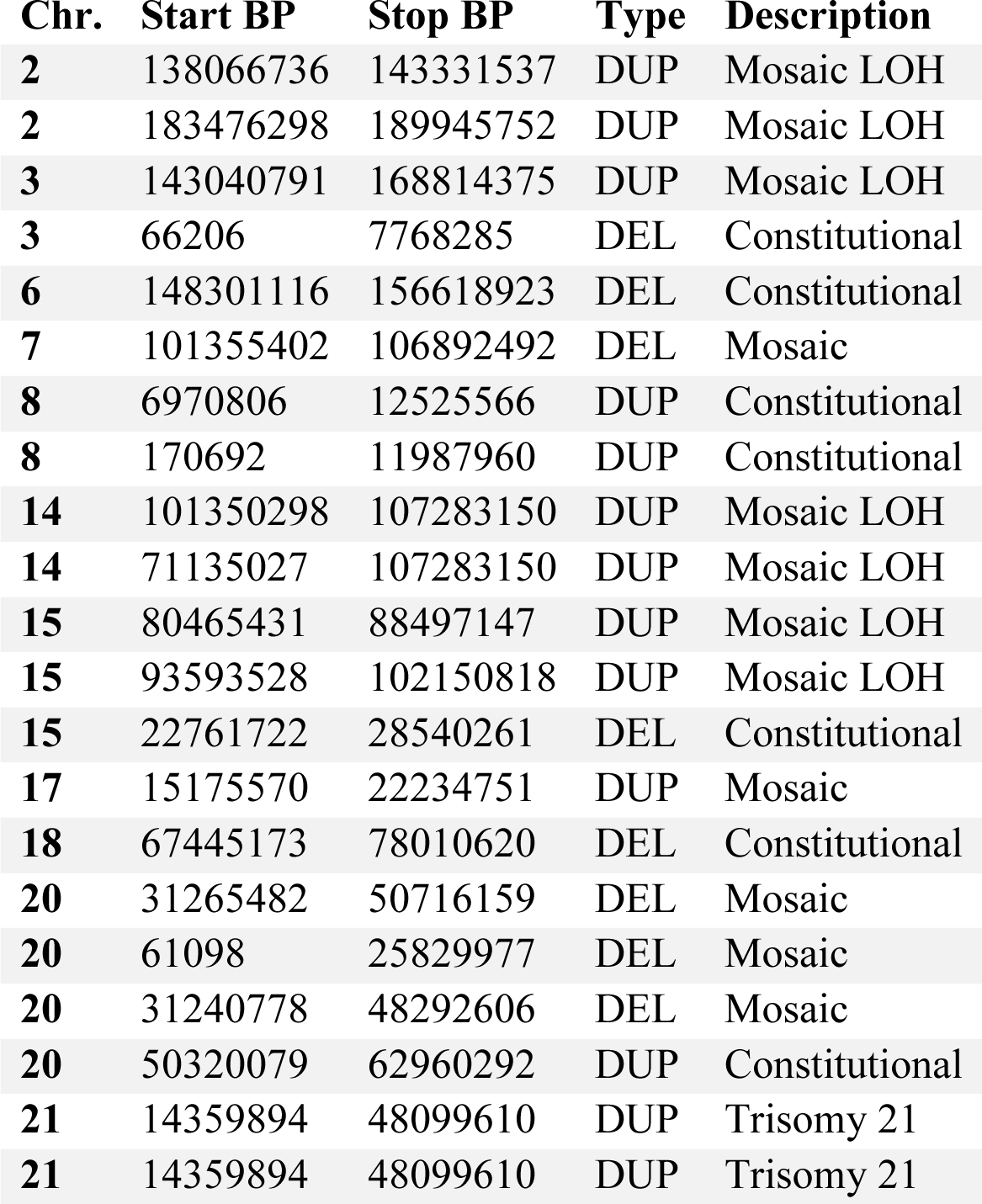
Large Genomic Events in BAVGWAS. Chr., Chromosome CNV on which CNV is located. Start BP, start base pair of CNV. Stop BP, stop base pair of CNV. Type, denotes if a CNV is a duplication (DUP) or deletion (DEL) event. Description, denotes if the CNV was a mosaic loss of heterozygosity (Mosaic LOH), loss of heterozygosity (LOH), mosaic (Mosaic), constitutional (constitutional), or trisomy 21 (Trisomy 21) event.

**S9 Table.**
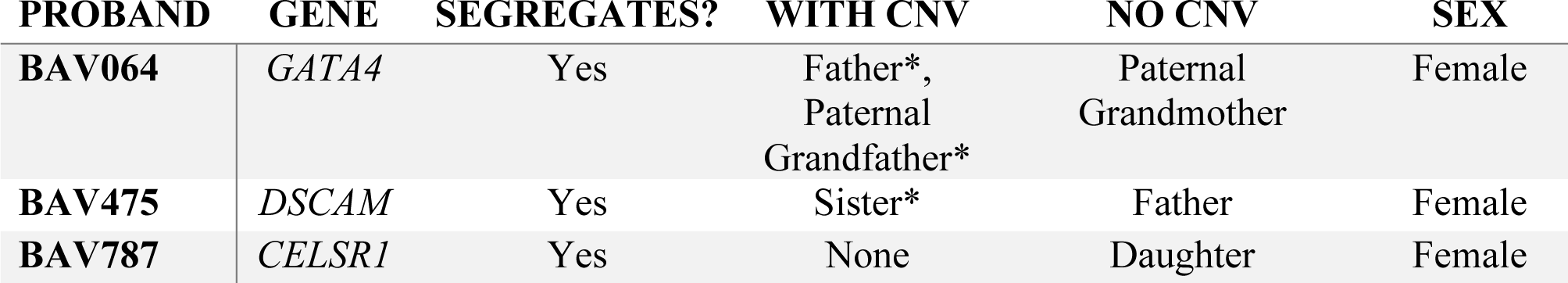
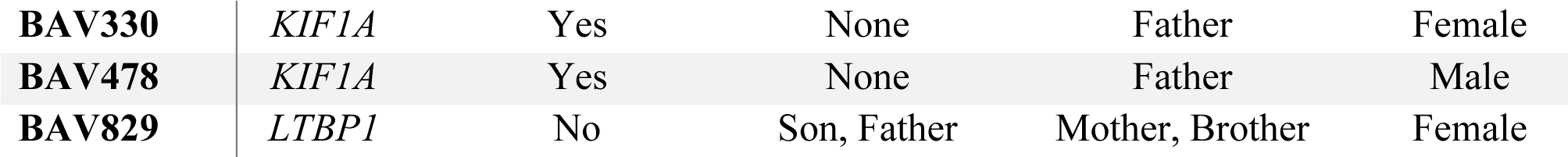
Pedigree Information for CNVs that Segregated with Disease. Proband, identification number of proband with CNV intersecting with gene of interest. Gene, gene of interest intersected by CNV. Segregates?, indicates if the CNV segregated with disease. Family With CNV, family members of proband that were found to have a CNV intersecting with the respective gene. Family Without CNV, family members of proband who were not found to have a CNV intersecting with the respective gene. Family members are listed if their genotype was available for the study. Sex, sex of the proband. *Indicates family members who also have BAV.

